# The role of prison-based interventions for hepatitis C virus (HCV) micro-elimination among people who inject drugs in Montréal, Canada

**DOI:** 10.1101/19008409

**Authors:** Arnaud Godin, Nadine Kronfli, Joseph Cox, Michel Alary, Mathieu Maheu-Giroux

## Abstract

**Background:** In Canada, hepatitis C virus (HCV) transmission primarily occurs among people who inject drugs (PWID) and people with experience in the prison system bare a disproportionate HCV burden. These overlapping groups of individuals have been identified as a priority populations for HCV micro-elimination in Canada, a country currently not on track to achieve its elimination targets. Considering the missed opportunities to intervene in provincial prisons, this study aims to estimate the population-level impact of prison-based interventions and post-release risk reduction strategies on HCV transmission among PWID in high HCV-burdened Canadian city, Montréal.

**Methods:** A dynamic HCV transmission model among PWID was developed and calibrated to community and prison bio-behavioural surveys in Montréal. The, the relative impact of prison-based testing and treatment or post-release linkage to care, alone or in combination with risk reduction strategies, was estimated from 2018 to 2030, and compared to counterfactual status quo scenario.

**Results:** Testing and linkage to care interventions implemented over 2018-30 could lead to the greatest declines in prevalence (23%; 95% Credible interval(CrI):17–31%), incidence (20%; 95%CrI: 10–28%), and prevent the most new chronic infections (8%; 95%CrI: 4–11%). Testing and treatment in prison could decrease prevalence, incidence, and fraction of prevented new chronic infections. Combining test and linkage to care with risk reduction measures could further its epidemiological impact, preventing 10% (95%CrI: 5–16%) of new chronic infections. When implemented concomitantly with community-based treatment scale-up, both prison-based interventions had synergistic effects, averting a higher fraction of new chronic infections.

**Conclusion:** Offering HCV testing and post-release linkage to care in provincial prisons, where incarcerations are frequent and sentences short, could change the course of the HCV epidemic in Montréal. Integration of post-release risk reduction measures and community-based treatment scale-up could also increase the impact of these interventions.

## 1. Introduction

Hepatitis C virus (HCV) is responsible for more years of life lost than any other infectious disease and could lead to a projected increase in the North American burden of end-stage liver disease and liver cancer over the coming years [1, 2]. The advent of highly efficacious and tolerable direct-acting antivirals (DAA) propelled the World Health Organization to set ambitious targets for HCV elimination by 2030 [3]. Micro-elimination, which consists of tailored interventions targeted at priority populations, was deemed a key approach to achieve this objective [3, 4]. Currently, Canada is not on track to attain these elimination targets, however, there is a national initiative advocating the adoption of micro-elimination strategies for people who inject drugs (PWID) and people in prison [5]. While the general Canadian population has a low HCV prevalence (0.96% in 2011), PWID and people in Québec’s provincial prisons, who have an average stay duration of 74 days per incarceration, experience a disproportionately high HCV burden, with antibody prevalence of 63% (2003-2015) for the former, and of 13% for the latter [6, 7, 8].

Provincial prisons have been identified as key settings to implement micro-elimination strategies. However, little efforts have been made to strengthen engagement along the HCV care cascade for people in Canadian provincial prisons [5, 9]. PWID have high incarceration rates in the provincial prison system and incarceration has been labelled as a driver of the epidemic [10]. Incarceration could sustain transmission through the elevated the risk of HCV transmission and acquisition in the period where formerly incarcerated PWID transition back to the community [10]. This heightened risk results from withdrawal symptoms experienced during incarceration and the unstable post-release environments, which can both lead to at-risk drug use behaviours in the community [11, 12, 13, 10, 14]. Tailored interventions for prevention, screening, and treatment, in prison or following release, could alleviate this heightened risk of onward HCV transmission [13, 9, 15]. PWID in Canada have negative and stigmatizing lived experiences in health care, which reduces their engagement in this system [16, 17]. Prisons represent important initial points of contact with these services [18, 16, 17]. Nonetheless, contrary to the Canadian federal prison system, there are no HCV-specific interventions in the country’s provincial prisons, with the exception of opt-out HCV testing in British-Colombia [9, 19].

Considering the missed opportunities to intervene in provincial prisons, a careful evaluation of potential interventions is necessary. One option is to allow people with chronic HCV to initiate treatment in prison. However, the majority of people in the provincial system serve sentences that are less than what is needed to complete a course of DAAs, which could lead to non-adherence or lower sustained virologic response (SVR) following release [20, 8, 9, 21]. Even if DAAs are completed and SVR is achieved during their stays, the heightened injecting risk post-release could lead to re-infection [22, 10, 9]. To alleviate such risks, interventions that enhance linkage to care with community services, such as nurse-led models could better address the needs of releasees [18, 23, 24]. Importantly, prison-based continuum of care interventions in provincial settings must be evaluated in the context of a broad DAA scale-up among community PWID, as advocated by the Canadian Blueprint for HCV elimination [5]. With the increased availability of pan-genotypic and short course DAA regimens, measuring the contribution of prison-based interventions to the overall micro-elimination effort is a current priority in Canada.

To address this, the current study aims to 1) assess whether a prison-based test-and-treat or a test and post-release linkage to care strategy is more effective in reducing HCV transmission, 2) quantify the added benefits of post-release risk-reduction measures to either approach, and 3) examine how a scale-up of DAA in the community modifies the effectiveness of these prison-based interventions. The population-level impact of the different scenarios will be measured among community PWID in Montréal, a high HCV prevalence setting in Canada. The results from this work will inform program managers and policy makers alike in the development of effective prison-based strategies contributing to the overall HCV elimination effort.

## 2. Methods

### 2.1. Model structure

A dynamic compartmental mathematical model of HCV transmission among PWID, adapted from Stone et al. [13], was parametrized, and calibrated to detailed longitudinal epidemiological data. The model is stratified by sex and considers three distinct but overlapping dynamics: 1) HCV transmission, 2) incarceration, and 3) injecting behaviours. The model is deterministic in nature and individuals without prior HCV exposure enter the population at a rate chosen to match PWID population size estimates from 2003 and 2010 [25]. Individuals either leave the model by all-cause or, when in advanced HCV disease stages, liver-related mortality. Active PWID and those recently released from prison have an increased risk of death as compared to individuals in the general population [26, 27]. A complete description of the model structure, the force of infection, and model equations are available in the supplementary appendix (Text S1).

#### 2.1.1. HCV transmission dynamics

PWID can acquire HCV and become acutely infected at a time-dependent force of infection that varies according to chronic prevalence, mixing patterns by incarceration history among community PWID, injecting behaviours, and interventions (i.e., needle and syringe programs; NSP) (Figure 1). Within six months, people with acute HCV either spontaneously clear their infection or develop chronic HCV. In the first case, people return to the HCV-susceptible stage, having developed HCV-specific antibodies, and can be re-infected. In the second case, people develop chronic HCV and progessive fibrosis until late HCV infection, where liver-related mortality can occur [28, 29]. People with chronic infection can be diagnosed at any disease stage and then treated at a time-varying rate that depends on fibrosis stage until 2018 (when fibrosis stage treatment restrictions were lifted in Canada) [30]. Treatment either leads to failure or SVR. Following SVR, people can be re-infected with HCV at the same force of infection as for primary infection [31, 32, 33].

**Figure 1:**
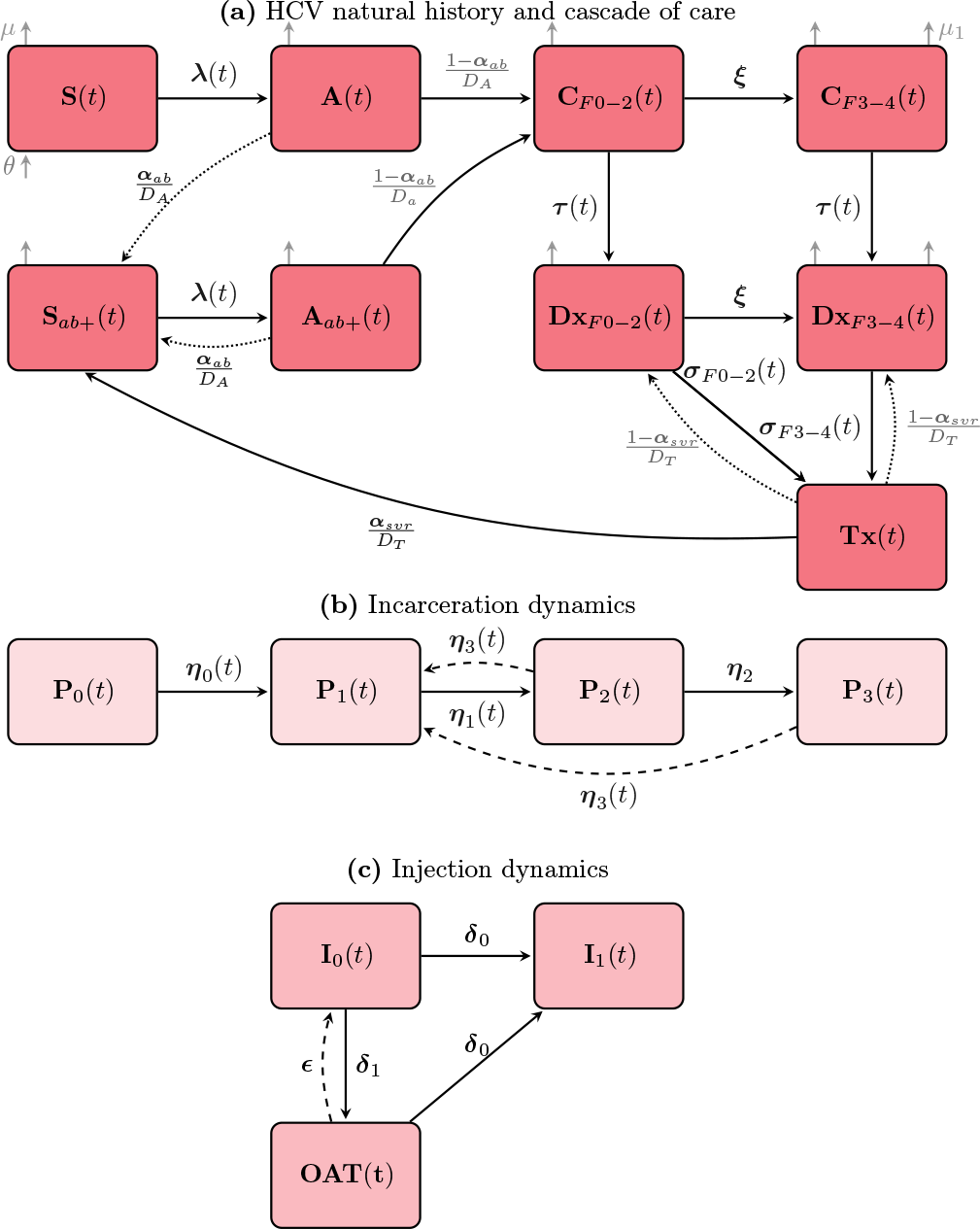
(a)Hepatitis C (HCV) natural history and cascade of care. The model is open, and people initiate injection as susceptible (**S**(*t*)) at a rate *θ*. Upon an effective contact, they become acutely infected (**A**(*t*)) at a time-dependent force of infection ***λ***(*t*). People spontaneously clear the infection after six months at a probability *α* and become susceptible but antibody postitive (**S**_*ab*+_(*t*)). Otherwise, they become chronically infected (**C**_*F* 0*−*2_(*t*)) and progess in fibrosis stages until late HCV infection (**C**_*F* 3*−*4_(*t*)), where they can die of liver-related mortality *µ*_1_. Chronically infected people can be diagnosed (**Dx**_*F* 0*−*2_(*t*), **Dx**_*F* 3*−* 4_(*t*)) without regards to disease stage at a rate ***τ*** and then linked to treatment (**Tx**(*t*)) at a time varying rate that depends on fibrosis stage (***σ***_*F* 0*−* 2_(*t*), ***σ***_*F* 3*−* 4_(*t*)). Treatment either leads to failure or sustained viral response and people become susceptibles but antibody positive. People who spontaneously cleared or were cured of the disease are susceptible to reinfection (**A**_*ab*+_) with HCV at the same force of infection ***λ***(*t*). (b) Incarceration dynamics. People who initiate injection have never been incarcerated (**P**_0_(*t*)) and can be incarcerated at a time-dependent rate ***η***_0_(*t*). They are then released back to the community at a time-dependent rate ***η***_1_(*t*) and are considered recently released for 6 months (**P**_2_(*t*)) after which they become previously released (**P**_3_(*t*)). People with experience in the prison system can be reincarcerated at a rate ***η***_3_(*t*). (c) Injection dynamics. People who inject drugs (PWID, **I**_0_(*t*)) completely stop injecting at a rate which is defined as the inverse of the average injecting duration (***δ***_0_). They can also initiate opioid agonist therapy (OAT) at a constant rate ***δ***_1_. On OAT, people can continue injecting and only stop after the average duration of injection ***δ***_0_. Once people have stopped they cannot go back to injecting.

#### 2.1.2. Incarceration dynamics

People who initiate injection are assumed to have no experience with the prison system and can be incarcerated at a time-dependent rate (Figure 1). At the end of their incarceration PWID are released back in the community and experience a short six-month period characterized by an increased risk of HCV acquisition and transmission, as well as heightened all-cause mortality [27, 10]. After this time, they are categorized as previously incarcerated [34, 10]. People released from prison and people with previous incarceration experience higher incarceration rates than that of primary incarceration [8, 35, 13].

#### 2.1.3. Injecting behaviours dynamics

PWID can completely stop injecting drugs, and a fraction can initiate opioid agonist therapy (OAT) to account for people in situations of opioid use disorder. On OAT, people can continue injecting, but with a lower risk of HCV acquisition and transmission and stop after an average active injecting duration determined from the literature [36, 37]. For simplicity, people who have completely stopped injection drug use (IDU) cannot re-initiate that behaviour (Figure 1).

### 2.2. Parametrization and calibration

#### 2.2.1. Parametrization

Two main data sources were used to inform our model parameters: repeated cross-sectional bio-behavioural surveys of PWID in Montréal (SurvUDI, 2003-2015) and two large prison bio-behavioral surveys conducted in 7 of the 17 provincial prisons in Québec (in 2003 and 2014) [7, 35, 8]. Parameters that could not be estimated from these local surveys were obtained from the relevant scientific literature. These include HCV-related biological parameters (spontaneous clearance, fibrosis progression rates, etc.), and relative risk measures for HCV acquisition and transmission, and death (Table 1). Where data was not available from meta-analyses, the most robust studies to inform model parameters were used (e.g., average injecting duration; Table 1). A detailed description of the abovementioned surveys is available in the supplementary information (Text S1).

**Table 1:**
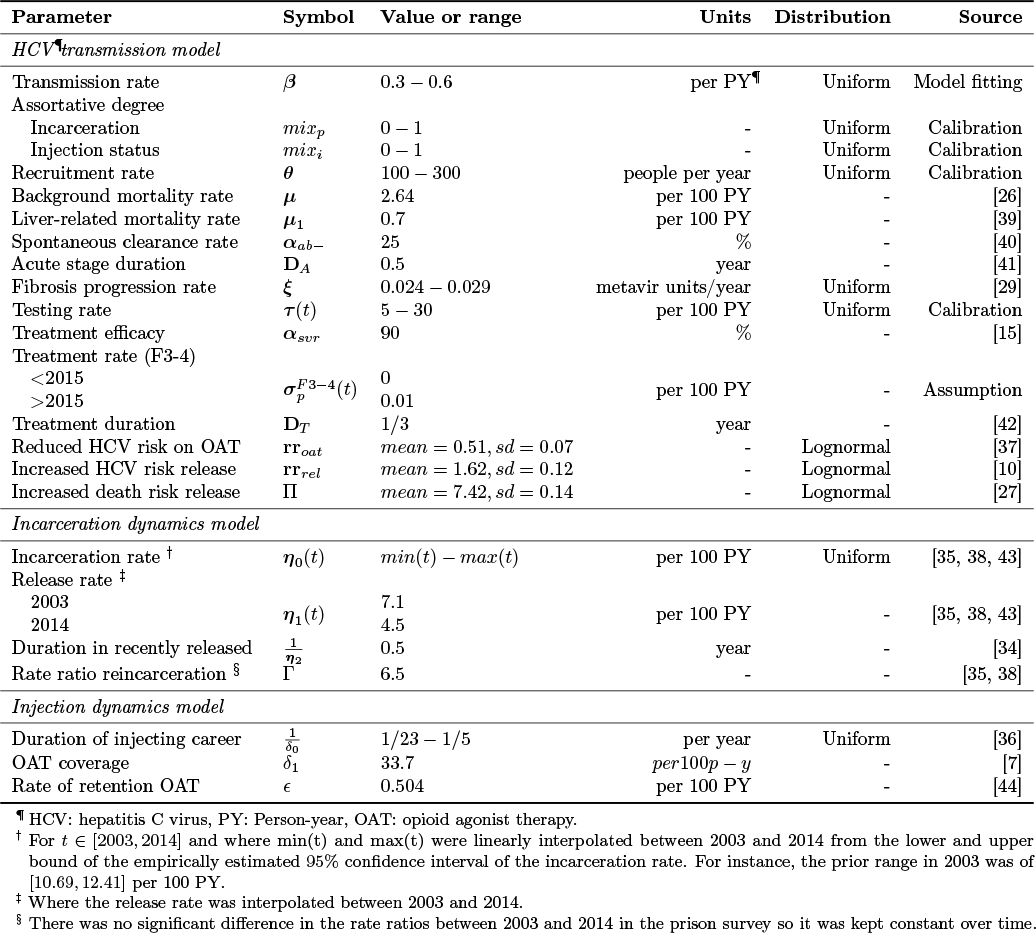
Description of model parameters, their values, prior distribution, and data sources.

#### 2.2.2. Calibration

The model was calibrated to annual HCV antibody prevalence and incidence estimates resulting from the SurvUDI network in Montréal (2003 to 2015), and antibody prevalence among incarcerated individuals self-reporting IDU in the past six months (in 2003 and 2014) [7, 38, 35]. Given its flexibility, a Bayesian framework was adopted for model calibration and appropriate prior distributions were elicited for relevant parameters (Table 1). Latin Hypercube sampling was used to sample 120,000 parameter sets from their prior distributions. After running the model to equilibrium in 2003, the posterior distribution of model outcomes was approximated with a sampling importance resampling algorithm, resulting in 1163 unique parameter sets (Text S2). Cross-validation of model predictions was also performed for knowledge of HCV status.

### 2.3. Intervention scenarios

Once calibrated to empirical data, we assessed the population-level impact of different scenarios on chronic HCV prevalence, incidence, the cumulative fraction of new chronic infections prevented, and the number of chronic infection averted per person treated over the 2018-2030 period, as compared to two couterfactual scenarios (Table 2). The first counterfactual kept testing and treatment rates as well as intervention coverage (i.e., OAT and NSP) at their 2018 levels. The second counterfactual scenario corresponds to an immediate community-based DAA scaleup, where community PWID are tested once per year and all diagnosed chronic cases systematically treated. Four prison-based intervention scenarios were implemented. In the first, 90% of people are tested and 75% of those diagnosed with chronic HCV are treated in prison (*PB T-Tx*).

**Table 2:** Description of the implemented scenarios in terms of HCV testing, treatment, and post-release risk reduction.

| Scenario | Testing | Treatment |  | Risk post-release |  |
| --- | --- | --- | --- | --- | --- |
|  |  | Prison | Post-release | HCV | Mortality |
|  | % <sup>a</sup> | % <sup>b</sup> | % <sup>c</sup> | - | - |
| Prison-based test-and-treat (*PB T-Tx*) | 90 | 75 | - | - | - |
| Post-release linkage to care (*PB T-L*) | 90 | - | 75 | - | - |
| *PB T-Tx* and risk reduction (*PB T-Tx + R*) | 90 | 75 | - | Halved | Halved |
| *PB T-L* and risk reduction (*PB T-L + R*) | 95 | - | 75 | Halved | Halved |

<sup>a</sup> Percentage of people tested during their stay in prison.
<sup>b</sup> Percentage of people treated during their stay in prison.
<sup>c</sup> Percentage of people linked to care and treated upon release.

In the second, 90% of people are tested in prison and 75% of people diagnosed with chronic HCV are linked to care for treatment upon release (*PB T-L*). The remaining two scenarios are the previous ones evaluated with a hypothetical complementary measure reducing the heightened post-release risk of HCV by 50% (*PB T-Tx + R* and *PB T-L + R*).

### 2.4. Sensitivity analyses

To assess the sensitivity of model results, we examined the correlations between parameter sets and model outputs. Specifically, the importance of three main parameters was investigated: 1) the rate ratio for HCV acquisition and transmission post-release, 2) the rate ratio for re-incarceration, and 3) the degree of assortative mixing by incarceration history in the community among those never incarcerated, recently released, and previously incarcerated. The correlation was examined for all outcomes for the test and post-release linkage to care scenario (*PB T-L*).

The model was implemented in Python 3.6 and solved with a validated Runge-Kutta algorithm of the 4^th^ order from the SciPy module [45].

### 2.5. Ethics

Ethics approval for this study was obtained from the Research Ethics Board of McGill University (IRB Study Number: A06-E43-18A) [35, 8].

## 3. Results

### 3.1. Model calibration

The model reproduced longitudinal trends in HCV antibody prevalence and incidence estimates (2003-2014) from SurvUDI, repeated cross-sectional surveys with information on 6,591 community PWID in Montréal (Figure 2)[7]. The model also replicated HCV-antibody prevalence from the two surveys of people in prison declaring IDU in the past six months, although the model prediction for 2014 could be slightly overestimated (Figure 2). Posterior distributions of model parameters can be found in the supplementary information (Table S2-1). Cross-validation of model outputs with empirical estimates of knowledge of HCV status also suggested excellent fits (Figure S2-2).

**Figure 2:**
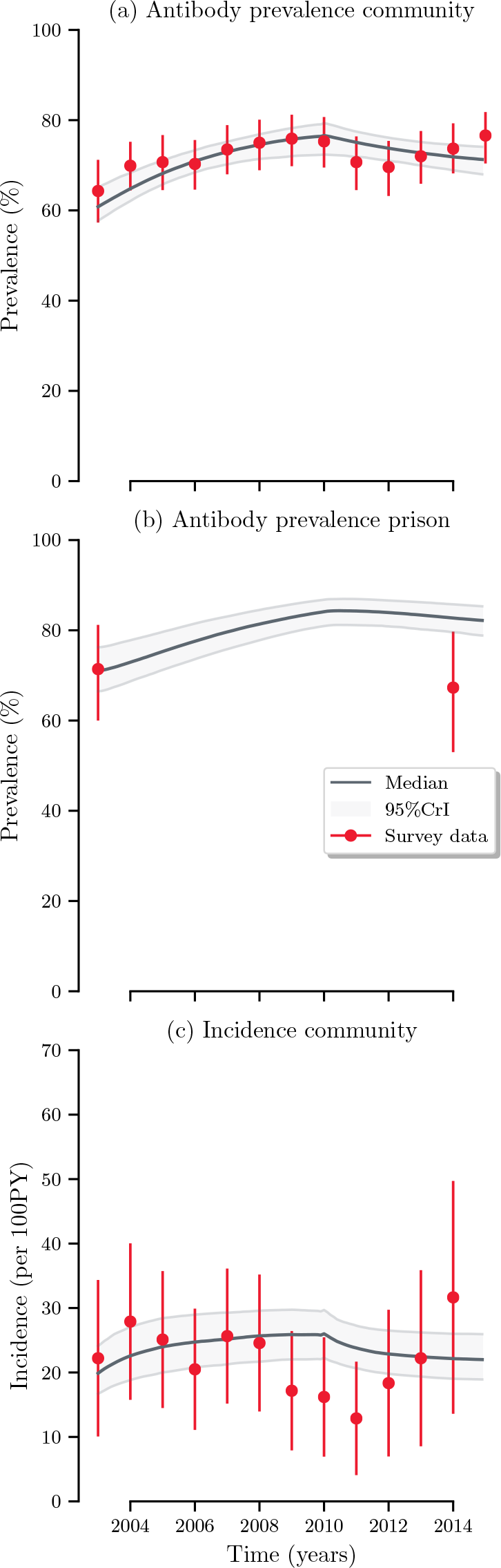
Model calibration to (a) antibody prevalence in the community, (b) in prison, and (c) to incidence in the community for 1163 unique parameter sets from 2003 to 2014. The empirical data points in (a) are estimates of antibody prevalence from the SurvUDI network among people at their first visit in the network. The empirical data points in (b) are estimates of antibody prevalence from the prison surveys among people who reported injecting drug use in the past six months. The empirical data points in (c) are estimates of incidence amongst repeaters from the SurvUDI network.

### 3.2. HCV epidemiology under the status quo

Maintaining the status quo, slight decreases in prevalence (8%; 95% Credible interval (CrI)(Cr): 4–10%) and incidence (5%; 95%CrI: 1–9%) are expected until 2030, which may result from a stable population with low community treatment, and harm reduction measures (OAT, NSP) already in place (Figures 3 and 4). The impact of all interventions on chronic HCV prevalence, incidence, prevented fractions, knowledge of HCV status, and fibrosis stages are presented in the supplementary information for 2030 (Table S2-2).

**Figure 3:**
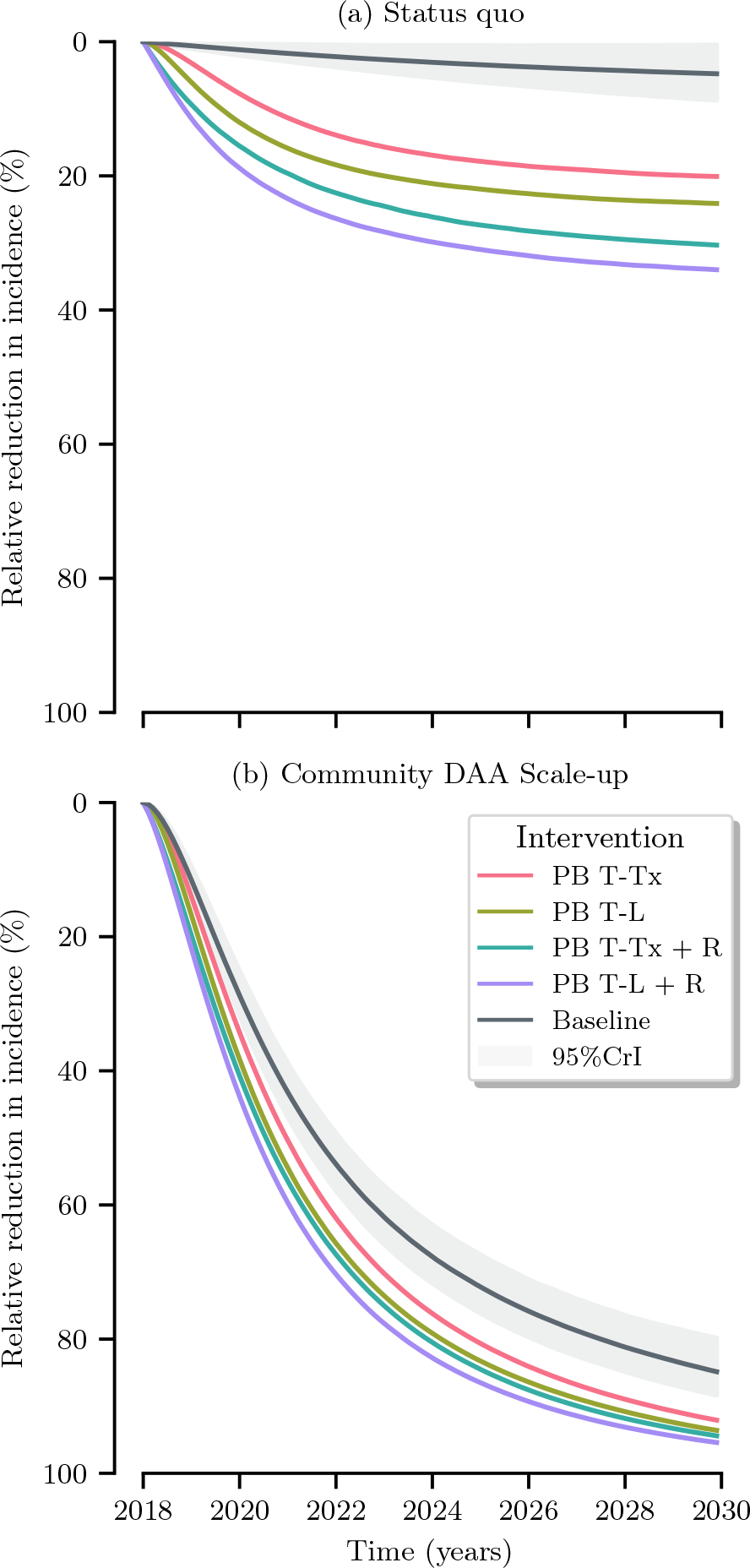
Relative reduction in incidence for interventions among active PWID in the community from 2018 to 2030 compared to (a) a counterfactual with no community scale-up of testing and DAA (b) a counterfactual with community scale-up of testing and DAA. Where *PB T-Tx* is a test-and-treat interventions in which 90% of people are tested and 75% of chronic cases are treated while in prison; *PB T-L* is a post-release linkage to care interventions where 90% of people are tested in prison and 75% are treated upon release; *PB T-Tx + R* and *PB T-L + R* are the previous interventions complemented by a post-release intervention that halves the risk of HCV acquisition and transmission.

**Figure 4:**
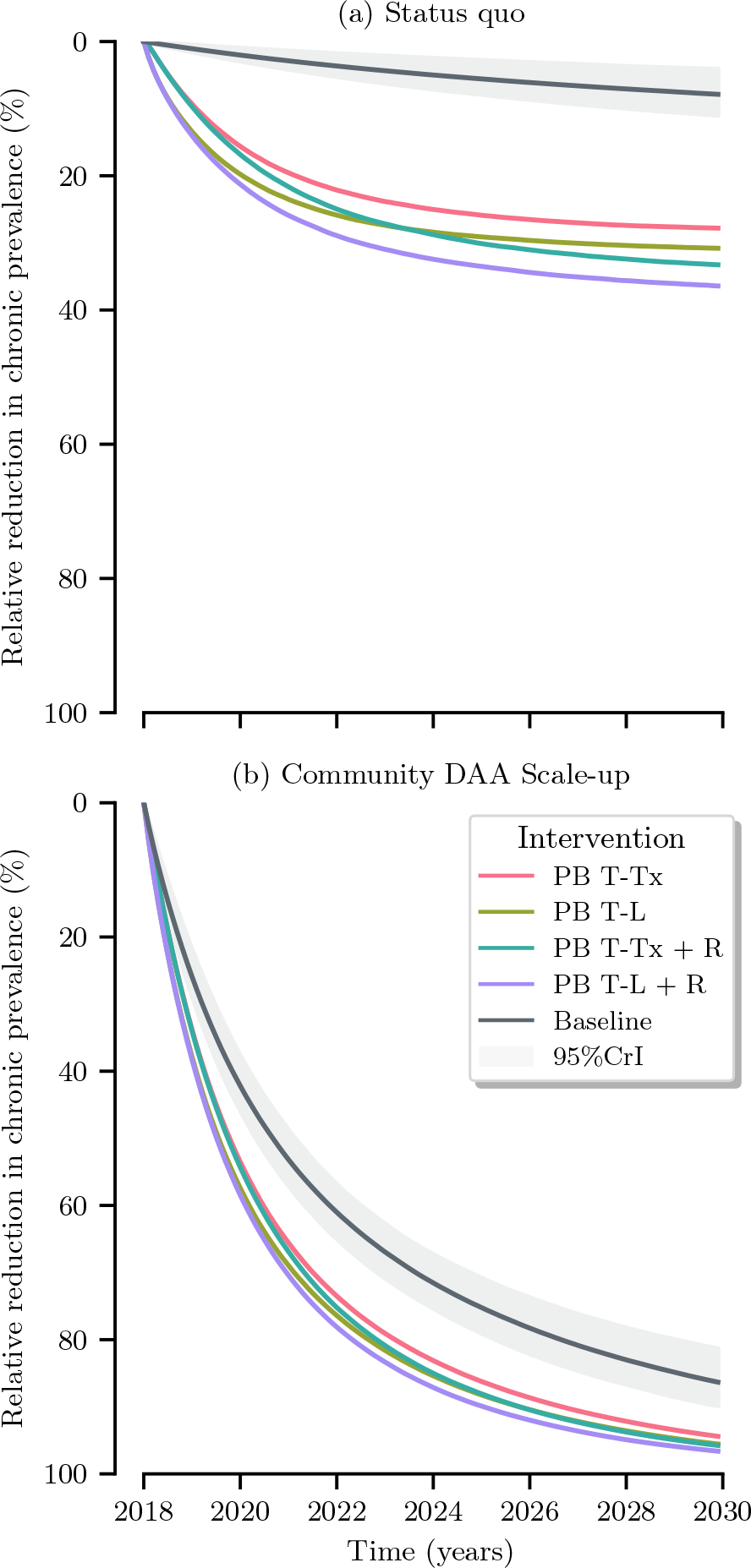
Relative reduction in chronic prevalence for interventions in active PWID in the community from 2018 to 2028 compared to (a) a counterfactual with no community scale-up of testing and DAA (b) a counterfactual with community scale-up of testing and DAA. Where *PB T-Tx* is a test-and-treat intervention in which 90% are tested and 75% of chronic cases are treated while in prison; *PB T-L* is a post-release linkage to care intervention where 90% of people are tested in prison and 75% are treated upon release; *PB T-Tx + Risk* and *PB T-L + Risk* are the same interventions where the elevated post-release risk is halved.

### 3.3. HCV epidemiology under a community DAA scale-up

If testing and treatment are scaled-up considerably in the community from 2018 onward, as part of micro-elimination efforts, substantial decreases in prevalence (86%; 95%CrI: 81–89%) and incidence (85%; 95%CrI: 79– 88%) could be expected by 2030 (Figures 3 and 4).

### 3.4. Impact of prison-based test-and-treat

A prison-based test-and-treat intervention (*PB T-Tx*) whereby 90% of people are tested and 75% of those diagnosed with chronic infection are treated during their prison stay would result in relative reductions of 27% (95%CrI: 20– 34%) in prevalence and 19% (95%CrI: 9–28%) for incidence as compared to 2018. The intervention could also prevent 7% (95%CrI: 20–34%) of new chronic infections over 2018-2030 (Figures 3-5), which represents 0.3 (95%CrI: 0.2–0.40) new chronic infections (including re-infection) averted per person treated (Figure 6).

**Figure 5:**
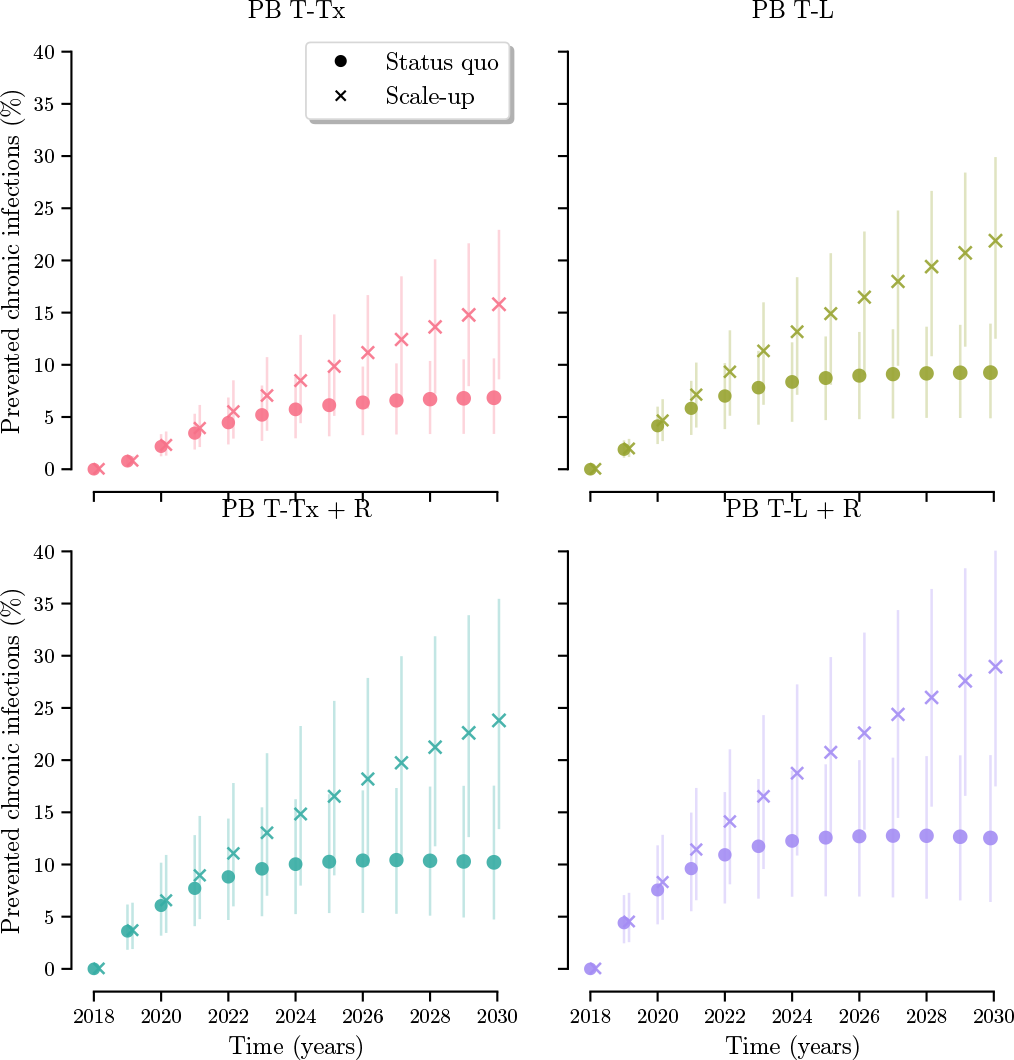
Prevented fractions of new chronic infections for all interventions among active PWID from 2018 to 2030 compared to the status quo and a community scale-up of testing and DAA. Where *PB T-Tx* is a test-and-treat intervention in which 90% are tested and 75% of chronic cases are treated while in prison; *PB T-L* is a post-release linkage to care intervention where 90% of people are tested in prison and 75% are treated upon release; *PB T-Tx + Risk* and *PB T-L + Risk* are the same interventions where the elevated post-release risk is halved.

**Figure 6:**
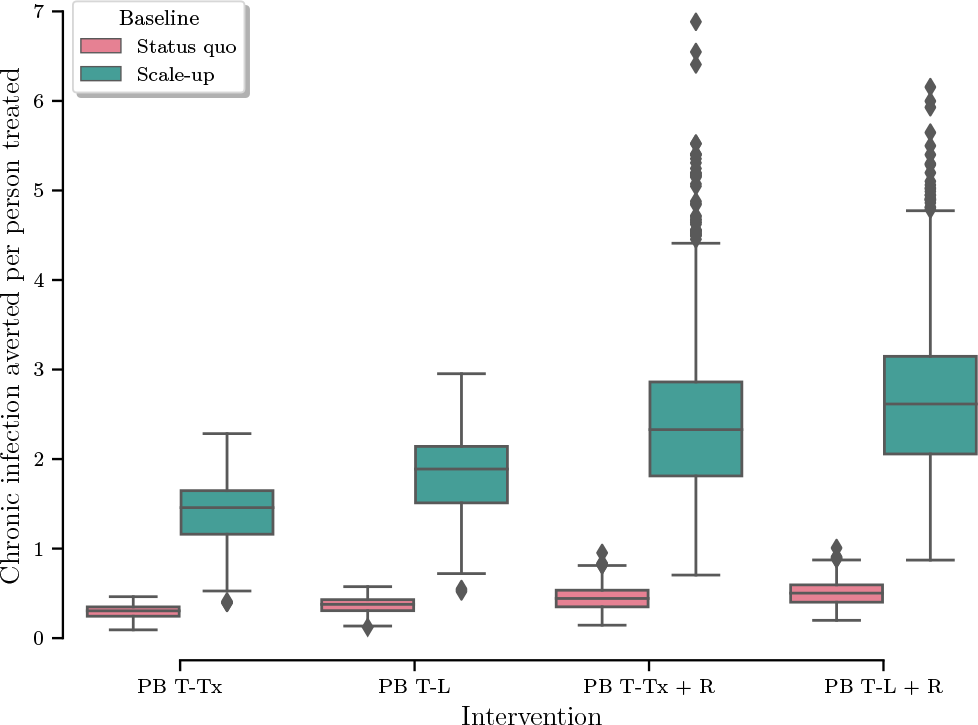
New chronic infections averted per person treated for all interventions in 2030 among active PWID compared to the status quo and a counterfactual with a community scale-up of testing and DAA. Where *PB T-Tx* is a test-and-treat intervention in which 90% are tested and 75% of chronic cases are treated while in prison; *PB T-L* is a post-release linkage to care intervention where 90% of people are tested in prison and 75% are treated upon release; *PB T-Tx + Risk* and *PB T-L + Risk* are the same interventions where the elevated post-release risk is halved.

The epidemiological impact of a prison-based test-and-treat strategy was magnified when combined with a community-based DAA scale-up (Figures 3 and 4). Treating 75% of people diagnosed with chronic HCV in prison prevented 14% (95%CrI: 8–20%) of new chronic infections after ten years, compared to community-based treatment scale-up alone (Figure 5). The prison-based intervention displayed synergistic effects with the community scale-up in terms of prevalence, incidence, and prevented fraction of new infections (Figures 3-5). As compared to a community-based scale-up alone, every additional person treated in prison averted 1.5 (95%CrI: 0.7–2.0) new chronic infections (Figure 6).

### 3.5. Impact of prison-based testing and linkage to care post-release

If 90% of incarcerated PWID are tested in prison and 75% of those diagnosed with chronic HCV are treated upon release (*PB T-L*), relative reductions of 30% (95%CrI: 22– 38%) for prevalence and of 23% (95%CrI: 11–33%) for incidence, as compared to 2018 levels, are expected by 2030 (Figures 3 and 4). Testing and post-release linkage to care also prevented 9% (95%CrI 5–14%) of new chronic infections compared to the status quo, and every additional person treated could avert 0.4 (95%CrI: 0.2–0.5) new chronic HCV infection (Figures 5 and 6).

Combining this testing and post-release linkage to care intervention with a community-based DAA scale-up led to a relative reduction in prevalence of 67% (95%CrI: 53–78%) compared to the scale-up alone by 2030. The intervention also lowered transmission, as compared to the community scale-up alone, by reducing incidence (57%; 95%CrI: 36– 70%) and preventing more new chronic infections (19%; 95%CrI: 11, 27%) (Figures 3-5). This combination also prevented more chronic infection per person treated compared to the community scale-up alone (1.9; 95%CrI: 0.9–2.6) (Figure 6).

### 3.6. Impact of prison-based interventions and post-release risk reduction

Both types of prison-based interventions had improved and sustained impacts when complemented by a hypothetical injecting risk reduction measures in the period following prison release. Prison-based test-and-treat (*PB T-Tx + R*) reduced prevalence and incidence by 32% (95%CrI: 25–41%) and 30% (95%CrI: 17–41%), respectively. As presented, the testing and post-release linkage to care intervention (*PB T-L + R*) achieved greater prevalence (36%; 95%CrI: 28–45%) and incidence (33%; 95%CrI: 20–45%) reductions, and prevented 13% (95%CrI: 7–20%) of new chronic infections by 2030 (Figures 3-5). Both interventions (i.e., *PB T-Tx + R* and *PB T-L + R*) could prevent slightly more new infections per person treated compared to their corresponding scenarios without a consideration of risk reduction measures, with 0.4 (95%CrI: 0.2–0.7) and 0.5 (95%CrI 0.3–0.7) additional chronic HCV averted per person treated for test-and-treat and testing and post-release linkage to care, respectively (Figure 6).

Testing 90% of people in prison and treating 75% of those identified with chronic HCV during their incarceration substantially reduced prevalence (68%; 95%CrI: 53– 81%) and incidence (62%; 95%CrI: 43–78%) until 2030, as compared to community-based DAA scale-up alone. The testing and linkage to care scenario had slightly increased reductions in prevalence (75%; 95%CrI: 61–85%) and incidence (69%; 95%CrI: 50–83%) compared to community-based DAA scale-up alone (Figures 3 and 4). Both prison-based interventions prevented numerous new infections, with 21% (95%CrI: 12, 32%]) for test-and-treat and 26% (95%CrI: 13–36%) for testing and post-release linkage to care. In the presence of a community-based DAA scale-up, 2.3 (95%CrI: 1.1–4.6) and 2.6 (95%CrI: 1.3–4.7) new chronic infections were averted for every additional person treated by 2030 for the test-and-treat and linkage to care scenarios, respectively (Figures 5 and 6).

### 3.7. Sensitivity analyses

Model results were primarily influenced by the degree of assortative mixing between different groups of PWID: those without experience with the prison system, those with recent release, and those with previous incarceration. As PWID mixed more within groups (and the degree of assortative mixing tended to 1), the relative impact of the scenario was lower. The increased incarceration rate did not influence model results much, nor did the rate ratio for the post-release risk of HCV acquisition and transmission (Fig. 7).

**Figure 7:**
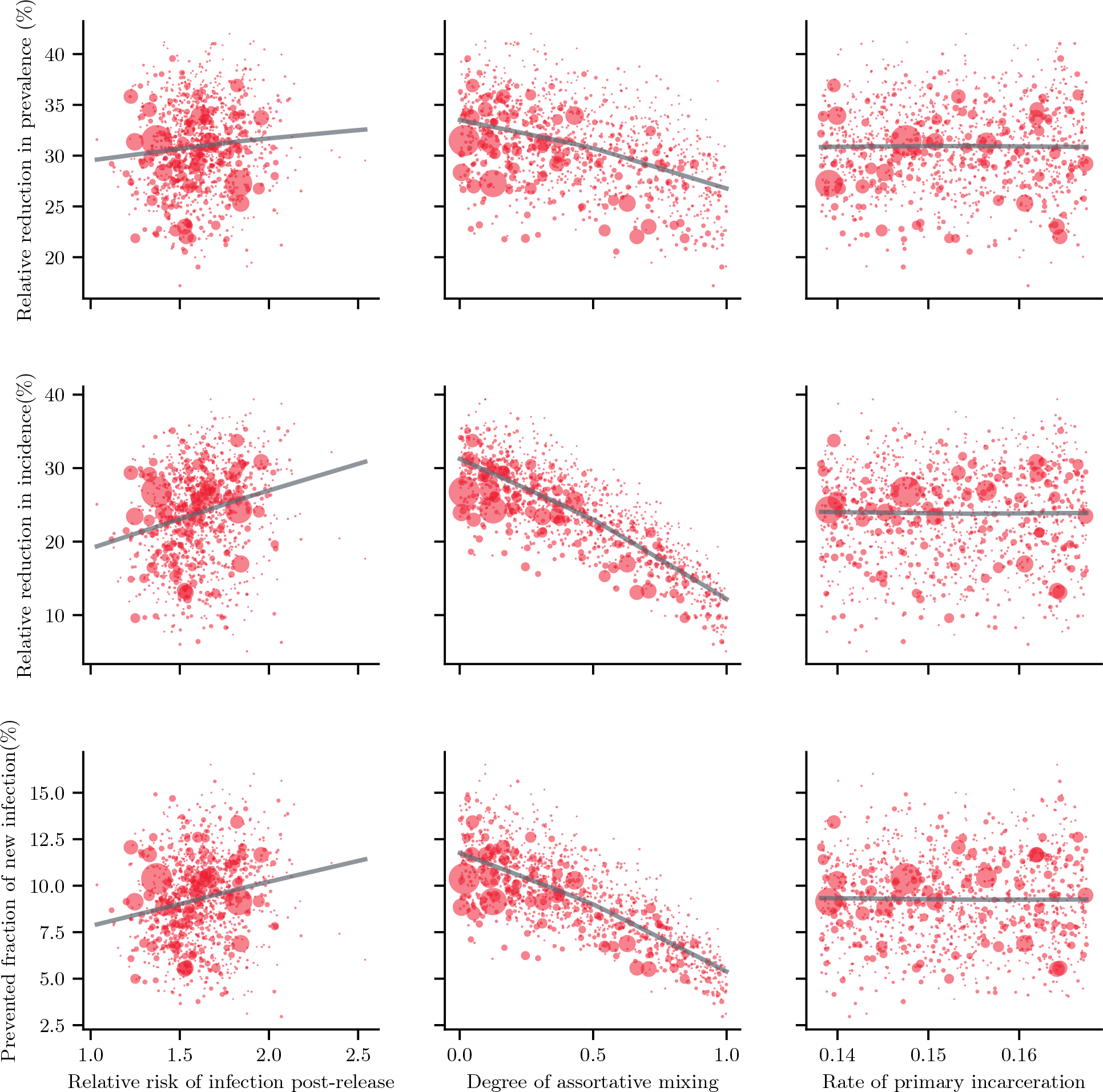
Sensitivity analysis for (from top to bottom) the relative reduction in prevalence, incidence and the prevented fraction of new chronic infections for (from left to right) the rate ratio for increased risk of HCV acquisition upon release from prison; mixing by incarceration status, where a degree of 1 means only assortative mixing (like-with-like); and the rate ratio for re-incarceration. The points represent parameter set and their size are relative to their resampling weight.

## 4. Discussion

HCV care in provincial prison settings, and the link between correctional and community services are currently inadequate [5]. Using a dynamic model of HCV transmission among PWID in Montréal, we showed that prison-based continuum of care interventions could play a central role in reducing HCV transmission among PWID. Importantly, the population-level impact of these interventions could be super-additive (i.e. more effective) when accompanied by a community-based DAA scale up. Overall, the results indicate that a multifaceted HCV response, strengthening community and prison care cascades, could be pivotal for launching efficient micro-elimination efforts.

In Québec’s provincial prisons, the sentence length per incarceration is very short [8, 20, 9]. This raised concerns regarding initiating treatment in prison, since lower SVR rates have been reported among inmates who are transferred to other prisons or released on treatment when compared to those who complete treatment in prison [21]. Test-and-treat interventions may have a greater impact in settings with longer duration of stay per incarceration, such as Scotland or federal prisons [13, 19]. In Montréal, interventions involving testing and post-release linkage to care could lead to larger relative decreases than prison-based treatment alone in HCV prevalence (30%) and incidence (23%) compared to the status quo, in part by decreasing the period during which people are susceptible to HCV re-infection (i.e., people on treatment cannot acquire HCV). Yet, the validity of this comparison hinges on achieving similar treatment rates in prison and post-release. The increased population health benefits of the linkage to care scenario could be compromised if higher treatment rates could be achieved inside prisons. One thing is clear, however, the current HCV care landscape misses out on in-carcerated PWID and people in provincial prison because interventions are not tailored to their needs.

The elevated post-release risk of HCV acquisition and transmission that occurs after release means that people can be rapidly re-infected with HCV, diminishing the effectiveness of prison-based HCV control measures. Breaking this instability cycle in the post-release period, which could promote drug use as a coping mechanism, is difficult [22, 46, 12]. Interventions that only seek to treat individuals are likely insufficient to sustainably reduce transmission if not complemented by risk-reduction measures in the post-release period [47]. Our results suggest, as found in Scotland [13], that integrated models of care in the post-release period could reduce the risk of HCV acquisition and transmission, and have the greatest epidemiological impact, preventing up to 26% of new chronic infections by 2030.

Although a hypothetical intervention reducing the post-release transmission risk by 50% was modelled in the present work, several real-world interventions could play this role and prevent re-infections.Nurse-led interventions in prison have helped increase treatment compliance, reduce risk behaviours, and manage disease burden: lessons learned from these successes could be applied for interventions in the post-release period [48, 49, 50]. Similarly, peer navigator programs could reduce the risk of IDU post-release and enhance engagement in the healthcare system by increasing trust, reducing perceived stigma, and improving social determinants of health, such as housing and employmentt [46, 51, 52].

Prison-based interventions may be important to curb ongoing transmission among PWID. These become even more critial in the context of a community-based DAA scale-up among PWID. In all investigated prison-based scenarios, treating one additional person could avert new chronic infections and thi impact was proportionally greater in the context of a community DAA scale-up. Treating people in prison or upon release allowed for a better HCV transmission control, mainly by reducing the risk of onward transmission in the post-release period [10, 12]. This result suggests that broad micro-elimination strategies for PWID should consider prison-based measures, as they target at-risk injecting risk periods more precisely and could use scarce financial resources more effectively.

Our results should be interpreted considering several limitations. First, and not unlike other studies, the data analyzed from the SurvUDI network and prison surveys is self-reported, and stigmatized injecting behaviours could have been underreported. For example, HCV prevalence from surveys in prison could be underestimated if recent infecting behaviours were underreported by participants interviewed inside prison walls. Second, even though uncertainty exists regarding available data on the PWID population size of Montréal, estimates suggest an important decline between 1996 and 2010, from 11,700 to 4,000 active PWID [25]. Without additional evidence post-2010 on demographic trends in the population injecting drugs, the size of the PWID population was kept stable from 2010 onward. Anecdotal evidence nevertheless suggests that it could be increasing. Third, the Montréal drug market has highly diversified in the past years and, while cocaine remains the main drug of choice, the use of crack and prescription opioids have increased [53, 7]. This could have led to changes in HCV incidence that were not mechanistically modelled, as drug type can impact injecting risk behaviours [54, 55, 53]. Fourth, for the hypothetical linkage to care scenario we chose a post-release linkage rate that is higher than that observed in recent studies [56, 57, 58] to reflect intensive micro-elimination efforts. Using lower linkage rates would unlikely change the qualitative interpretation of our findings, which compares the different interventions in relative terms. Nevertheless, this points to the potential lower impact of prison-based interventions if linkage and treatment rates cannot be reached and suggests that HCV elimination among PWID may only be achieved with a broad and intensive community-based scale-up of treatment. Finally, our results may only be generalizable to settings with similar polydrug use patterns, incarceration dynamics, and patient demographics.

There are several strengths to this study. First, we were able to replicate HCV epidemic trends in both community and prison, two settings that are often difficult to study simultaneously. Second, the data-rich environment of Montréal allowed for robust quantification of model parameters and inclusion of several calibration outcomes [7, 35, 38]. Third, a 12-week duration and an SVR rate of 90% were conservatively assumed for DAA therapy, with perfect adherence. However, new drugs with eight-week duration, assuring treatment completion and SVR, are currently being evaluated in multiple settings and could further enhance the ease of treating people with chronic HCV in prison or upon release [59, 60]. Last, parameter uncertainty was considered through Bayesian model calibration and sensitivity analyses were performed.

## 5. Conclusion

Sustainably curbing HCV transmission among PWID in Montréal will require the rapid scale-up of interventions both in prison and in community settings, and should leverage their synergistic impacts. Prison-based intervention strategies could have an important impact on the disease burden in this priority population. Enhancing linkage between community and prison healthcare services would notably be important in Québec’s provincial prison, where sentence lengths may be too short to complete treatment. Nonetheless, intervention programs comprising treatment in and upon release from prison, based on sentence length, should also be evaluated in future work. Furthermore, public health authorities should consider implementing risk reduction initiatives targeted at the short period soon after release from prison. Overall, the results presented above suggest that reaching the goal of HCV elimination among PWID in Montréal would be facilitated by improving integration and coordination of both correctional and public health services. In Canada, only three provinces (Nova Scotia, Alberta, and British Colombia) have integrated prison health care, including HCV, in the Ministry of Health’s mandate. Elsewhere, the ministry responsible for corrections oversees healthcare of people in prison [61]. Transitioning toward a more integrated system could prove challenging, however, and empirical studies should be conducted to assess acceptability, feasibility of such initiatives. Results from the current study can guide stakeholders to close the gaps between these two distinct settings, which will be crucial in the efforts to eliminate HCV but also in improving the health of a marginalized group of individuals with limited access and engagement in healthcare.

## Data Availability

Online supplemental material is available with specification on model structure and methods for model calibration. Data used for calibration is publicly available from published reports and studies.

## 7. CReDiT author statement

**Arnaud Godin:** Conceptualization, Methodology, Data curation, Formal Analysis, Software, Visualization, Writing – original draft. **Nadine Kronfli:** Conceptualization, Methodology, Validation, Writing – review & editing. **Joseph Cox:** Conceptualization, Methodology, Supervision, Writing – review & editing. **Michel Alary:** Investigation, Methodology, Supervision, Writing – review & editing. **Mathieu Maheu-Giroux:** Conceptualization, Methodology, Supervision, Validation, Resources, Writing – review & editing. All authors approved the final version of this manuscript.

## 8. Acknowledgement

The authors wish to acknowledge Charlotte Delaunay-Lanièce, Carla Doyle, Rachael Midwild, and Katia Giguère for their comments and suggestions in all phases of this project.

